# Paramagnetic rim lesions are associated with pathogenic CSF profiles and worse clinical outcomes in multiple sclerosis: a retrospective cross-sectional study

**DOI:** 10.1101/2022.01.08.22268838

**Authors:** Christopher C. Hemond, Jonggyu Baek, Carolina Ionete, Daniel S. Reich

## Abstract

**Background and Objective:** Paramagnetic rims have been observed as a feature of some MS lesions on susceptibility-sensitive MRI and indicate ongoing inflammation, principally consisting of compartmentalized activated microglia/macrophages. We investigated clinical, MRI, and intrathecal (cerebrospinal fluid, CSF) associations of paramagnetic rim lesions (PRL) using 3T MRI in MS.

**Methods:** This is a retrospective, cross-sectional analysis of patients at a single neuroimmunology clinic. All patients had standardized 3T MRI using a multiecho T2*-weighted sequence with susceptibility postprocessing (SWAN protocol, GE) as part of the inclusion criteria. SWAN-derived filtered phase maps and corresponding T2-FLAIR images were manually reviewed by one expert rater blinded to clinical data, and PRL were determined based on qualitative assessment of hypointense paramagnetic edges on corresponding T2-hyperintense lesions. Descriptive statistics, t-tests, ANOVA, and linear regression determined demographic, clinical, MRI, and intrathecal profile associations with the presence of one or more PRL.

**Results:** One hundred and forty-seven (147) MS patients were included in this analysis (2 clinically isolated syndrome, 118 relapsing-remitting, 14 secondary progressive, 13 primary progressive). Baseline mean age was 48.8 years, disease duration 12.8 years, and median EDSS 2, with 79% women. Seventy-five percent of patients were receiving a disease-modifying therapy, and 79 patients (54%) had available cerebrospinal fluid (CSF) analysis. Sixty-three patients (43%) had at least 1 PRL. PRL status (presence or absence) did not vary by sex or EDSS but was associated with younger age (51 vs 46 years; p=0.01) and shorter disease duration (14.5 vs 10.5 years; p=0.01). PRL status was also associated with worse disease (MS severity score: 2.8 vs 3.7; p=0.05) and blood-brain barrier disruption as determined by higher protein and pathologically elevated albumin quotient, as well as the presence of CSF oligoclonal bands (all p≤0.05); there was no association with immunoglobulin index or synthesis rate. PRL status was additionally associated with higher burden of T2-hyperintense cerebral lesion volume (T2LV), higher age-adjusted cerebral brain volume loss (especially of gray matter), and poorer performance on multiple clinical measures, including the 9-hole peg test and symbol digit modalities test (but not timed 25-foot walk speed). Clinical and intrathecal profiles remained associated with PRL after adjustment for age and in many cases T2LV as well. Sensitivity analyses limited to subgroups of patients without disease activity at the time of CSF sampling remained supportive of results. Patients with PRL were being treated with higher-efficacy disease-modifying therapies at the time of the data query.

**Conclusions:** PRL, an emerging noninvasive biomarker of chronic cerebral neuroinflammation in MS, are confirmed to be associated with greater disease severity and newly shown to be associated with intrathecal inflammation and blood-brain-barrier disruption.

## Introduction

Multiple sclerosis (MS) is a chronic, inflammatory neurological disease most often characterized by recurrent episodes of immune-mediated demyelination of the central nervous system. Most patients eventually suffer concurrent neurodegeneration, characterized indirectly as accelerated brain and spinal cord tissue loss compared to age-matched healthy populations and progression of clinical symptoms in the absence of new focal inflammatory lesions. The mechanisms of this neurodegenerative process are likely multifactorial; one component is thought to be related in part to chronic, compartmentalized CNS inflammation [1]. Histological (postmortem) analysis has revealed immunologically activated, iron-enriched microglia at the edges of a subset of chronically active MS lesions; this sequestered iron is visible as a paramagnetic rim on susceptibility-sensitive MRI sequences [2, 3]. These paramagnetic rim lesions (PRL), as assessed on 3T MRI, have been recently shown to correlate with higher neurological disability and disease severity, lower brain volumes, and longitudinal clinical deterioration [4, 5]. The cerebrospinal fluid profile of PRL has not yet been characterized. Here we aimed to evaluate the intrathecal profile in MS with the hypothesis that PRL would be associated with signs of higher chronic intrathecal inflammation.

## Methods

### Consent to Participate

This study was reviewed and approved by the University of Massachusetts ethics board (IRB Protocols # H00016906 and 14143). Data collection, storage, and access were in accordance with the Health Insurance Portability and Accountability Act.

### Study Design and Subjects

This is a retrospective analysis of patients at a single neuroimmunology clinic located at a large academic medical center. All subjects were participating in at least one of two ongoing prospective, longitudinal observational studies. Inclusion criteria were having a confirmed diagnosis of a clinically isolated syndrome or MS based on 2010 McDonald criteria, age <75, and at least one MRI performed on the same 3T MRI scanner with harmonized MS imaging protocol as detailed below. Baseline was considered as the time of the first eligible MRI, with clinical and laboratory measures calculated relative to that value. Patient neurological disability ratings, disease-modifying therapy use, and most laboratory data were obtained by manual chart review from the electronic medical record. Some laboratory and clinical data were also obtained via electronic query. Clinical and laboratory data were similarly chosen based on proximity to the time of the baseline MRI. All EDSS scores and MS diagnoses/phenotypes extracted from the chart were rated by expert clinical neurologists with dedicated subspecialty training in neuroimmunology and MS. Clinical assessments (timed 25-foot walk, symbol-digit modalities test, and modified 9-hole peg test – dominant hand only) were performed by trained medical staff. The Multiple Sclerosis Severity Score (MSSS) was calculated based on tabulated values [6].

All CSF-related data were manually recorded from the medical record. For CSF-leukocyte values, tube 4 of the CSF collection was used for assessment. Albumin quotient was calculated as the ratio of CSF albumin to serum albumin multiplied by 1000. Pathological cutoffs of albumin quotient were defined by an age-adjusted equation: age/15 + 4 [7]. If CSF was obtained on more than one occasion, the data closest to the time of MRI were used. Two categories were defined for determination of disease activity near the time of the lumbar puncture (LP). First, manual review of the electronic medical record was performed; any clinical MS flares that occurred in the 3 months period prior to LP — as determined by expert clinician documentation and/or the administration of high-dose IV methylprednisolone — were recorded. Radiology reports of any MRI scans were reviewed in the 3 months prior to LP, and any gadolinium-enhancing or new T2-hyperintense lesion (if available) reported in the brain or spinal cord were recorded. Three-month disease activity was dichotomously defined as any patients having evidence of either clinical or MRI disease activity within the 3-month period prior to LP. A second measure of disease activity was defined as any patient with one or more gadolinium-enhancing lesion of the brain or spinal cord within the 1-month period before or after LP. The LP setting (inpatient vs outpatient) and reason (diagnostic or otherwise) was also recorded from the clinical documentation.

### Image Acquisition

All assessed images were acquired on a 3T MRI scanner (Signa Pioneer, General Electric Healthcare, Wisconsin, USA) using a 32-channel head and neck coil and a consistent acquisition protocol for MS patients. Pre-gadolinium 3D sagittal T1-weighted images were acquired with the BRAin VOlume (BRAVO) Fast Gradient Echo sequence as follows: TR/TE/flip angle = 7.5ms/3.0ms/8°, nominal field of view=240mm, frequency/phase=220/220, slice thickness=1.2, reconstructed voxel size = 1.20×0.94×0.94mm, scan duration=3:37; sagittal 3D T2-FLAIR (FLuid Attenuated Inversion Recovery): TR/TE/flip angle/echo train length = 5400/133/90°/140, nominal field of view=250mm, frequency/phase=256/224, slice thickness=0.8 at 50% resolution, reconstructed voxel size=0.80×0.49×0.49mm, scan duration=3:34; 3D susceptibility-sensitive imaging (Susceptibility Weighted ANgiography, i.e. SWAN protocol): TR/TE/flip angle/echo train length = 42ms/24ms/10°/4, slice thickness/overlap = 3.0mm/1.5mm, reconstructed voxel size=0.47×0.47×3.0mm, scan duration=2:27. If gadolinium was administered, an additional sagittal 3D T1-weighted fast spin echo (CUBE, no acronym) scan was obtained approximately 10 minutes after the intravenous infusion of gadoterate meglumine with the following parameters: TR/TE/flip angle 435ms/16.1ms/90°, nominal field of view=250mm, frequency/phase=256/256, slice thickness=0.6, reconstructed voxel size = 0.60×0.49×0.49mm, scan duration=4:36.

### Image Analysis

MRI data were reconstructed using manufacturer software on the scanner and exported in DICOM format, from which they were converted to “NIFTI” format using “dcm2niix” software (version 1.0.20201102) [8]. For each subject, multimodal within-subject MRI sequences were visualized and coregistered as needed (mutual information criteria; rigid transformation with 6 degrees of freedom) in the software ITK-SNAP [9]. Manufacturer-reconstructed Susceptibility-Weighted ANgiography (SWAN) filtered phase images (“right hand” convention, paramagnetic = dark), as well as the T2-FLAIR, pre-gadolinium T1-weighted, and post-gadolinium (if administered) T1-weighted sequences, were then simultaneously reviewed in the axial plane for each session by a single investigator (C.H., a neurologist with more than 5 years of subspecialty training and experience with MS neuroimaging) initially blind to clinical data. This reviewer identified MS lesions >3mm in largest diameter [10] on the T2-FLAIR sequence and compared these to the coregistered, filtered-phase SWAN and T1-weighted images. Confluent periventricular lesion complexes were included for assessment when distinctly identifiable lesions could be visualized perpendicular to the periventricular complex. PRL were defined using the filtered-phase SWAN as having distinct hypointense edges that correspond with at least 50% of the T2-FLAIR lesion perimeter, as well as an isointense central area, as defined by prior investigators [11]. Qualitative T1 characteristics (hypointense, isointense) and lesional contrast enhancement (if administered) were additionally recorded. Any lesions that demonstrated contrast enhancement on T1-weighted images were excluded, since they may have transient paramagnetic rims that do not necessarily indicate the presence of chronic neuroinflammation [12].

### Quantitative MRI Analysis

All T1-weighted scans were manually reviewed for image quality to assure no significant artifacts. Images were then analyzed using the SIENAX pipeline, part of the FMRIB software library (FSL, v5.0) customized with dedicated brain extraction using the OptiBet software [13]. This SIENAX analysis yielded segmented volume data for the white matter, gray matter, and whole brain, with a normalization multiplicative based on skull registration to a template. FSL-FIRST was used to segment and obtain subcortical gray matter volumes, including of the thalamus. T2-hyperintense lesion number and summative volumes (T2LV) were segmented from T2-FLAIR images using the lesion prediction algorithm [14] as implemented in the LST toolbox version 3.0.0 (www.statistical-modelling.de/lst) for the Statistical Parametric Mapping toolbox (https://www.fil.ion.ucl.ac.uk/spm) with manual adjustment as needed for gross errors in ITK-SNAP.

### Statistical Analysis

Normality of variables was assessed by a combination of skew/kurtosis metrics and visual inspection with the histogram, as needed. Chi-square, ANOVA, or nonparametric (Mann-Whitney or Kruskal-Wallis) tests were used for comparison of PRL category with clinical, MRI, and laboratory data. Linear regression with robust standard errors was used for univariable and age-adjusted multivariable analyses, with PRL included as a dichotomous or ordinal variable for comparison with prior studies [4, 15] and to determine “dose-dependent” effects. Additional multivariable models were explored as sensitivity analyses. All statistical analyses were performed in Stata v14.

## Results

### Cohort description

One hundred and forty-seven patients were included, after the exclusion of 6 who had severely degraded MRI quality (n=5) or no T1-weighted image (n=1). The mean age ±SD of the cohort was 48.8±11.7 years; 120 were relapsing phenotype (118 RRMS, 2 CIS), and 27 were progressive (14 SPMS; 13 PPMS); 77% were women. These details, as well as clinical characteristics and comparisons between the subgroup of patients with available CSF, are summarized in Table 1. Notably, the subgroup with available CSF results was characterized by shorter disease duration (14.8±9.0 versus 11.0±10.3 years, p=0.02), greater disease severity (MSSS mean 2.7±2.5 versus 3.6±2.3 years, p=0.04), and more likely to be on higher-efficacy therapy (χ^2^ =12.5, p=0.01). Eighty-one scans (55%) were administered with gadolinium contrast; 10 (12%) of these patients had at least one gadolinium-enhancing lesion (mean 1.8±1.2, range 1-4). CSF was obtained on average 3.8±4.0 years prior to MRI assessment. The reason for performing lumbar puncture were predominantly diagnostic (n=61, 77%), but included cases of assessment for the JC virus on high-risk therapies (n=18, 23%) as well. Most lumbar punctures were performed in the outpatient setting (n=58, 73%) compared to inpatient (n=21, 27%). In the 3 months prior to lumbar puncture (LP), 36/79 (46%) of patients had evidence of clinical or MRI disease activity. Of those who had an MRI with contrast within 1 month of the LP (n=52), 21 (40%) had at least one gadolinium-enhancing lesion (Table 2).

**Table 1:**
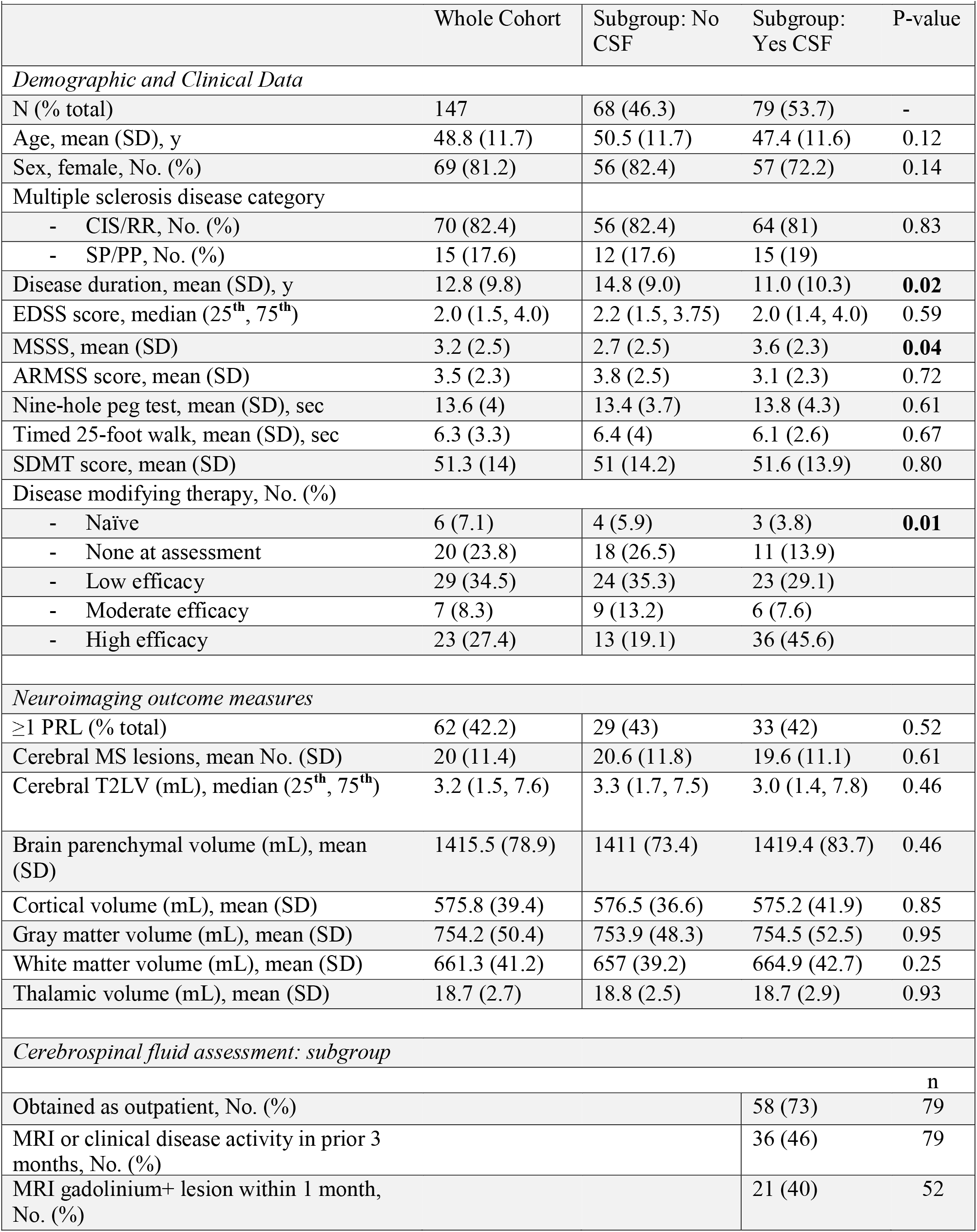
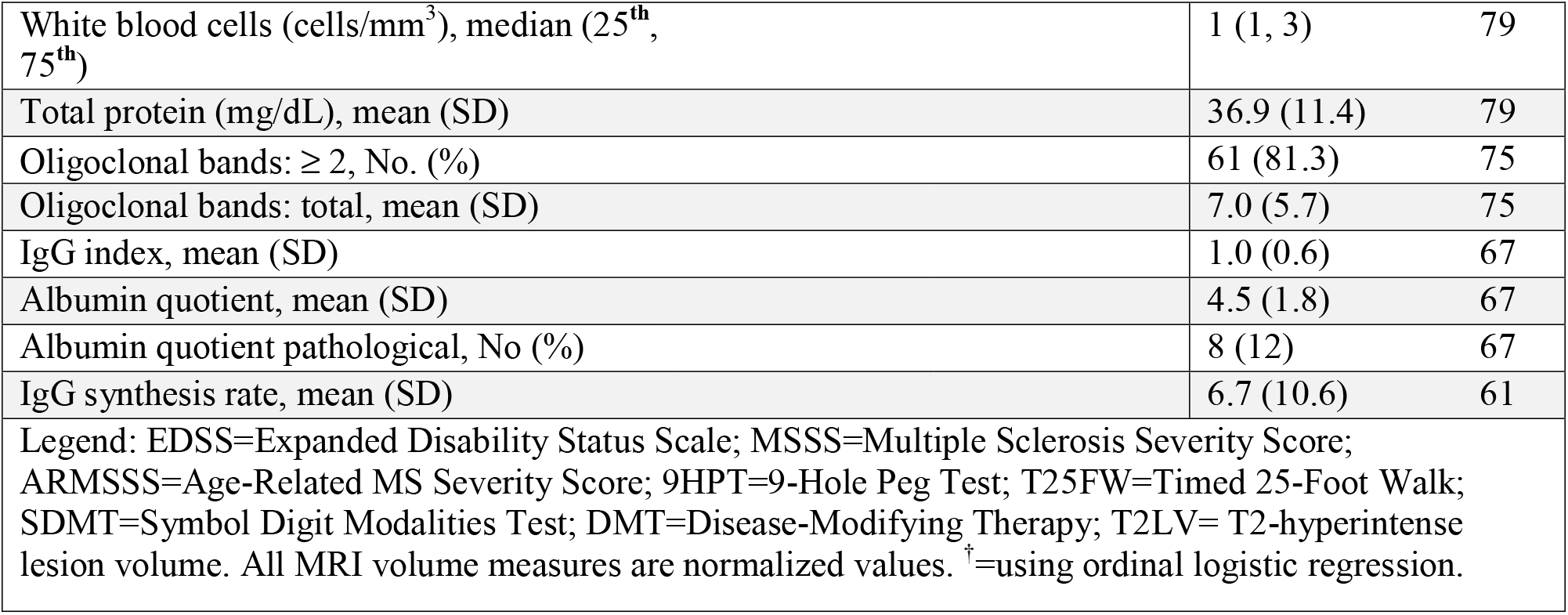
Summarized demographic, clinical, laboratory and MRI characteristics of the cohort

**Table 2:**
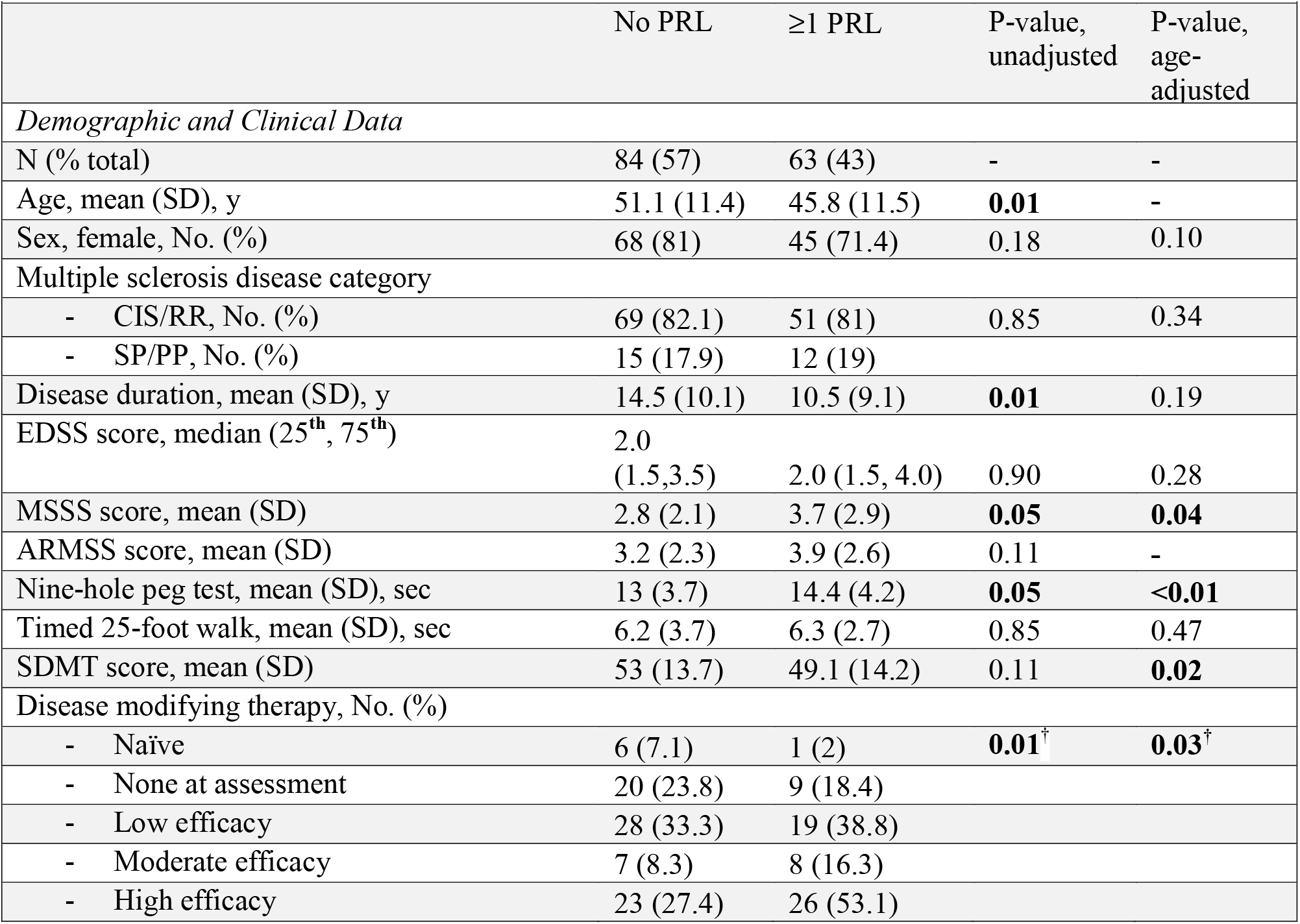
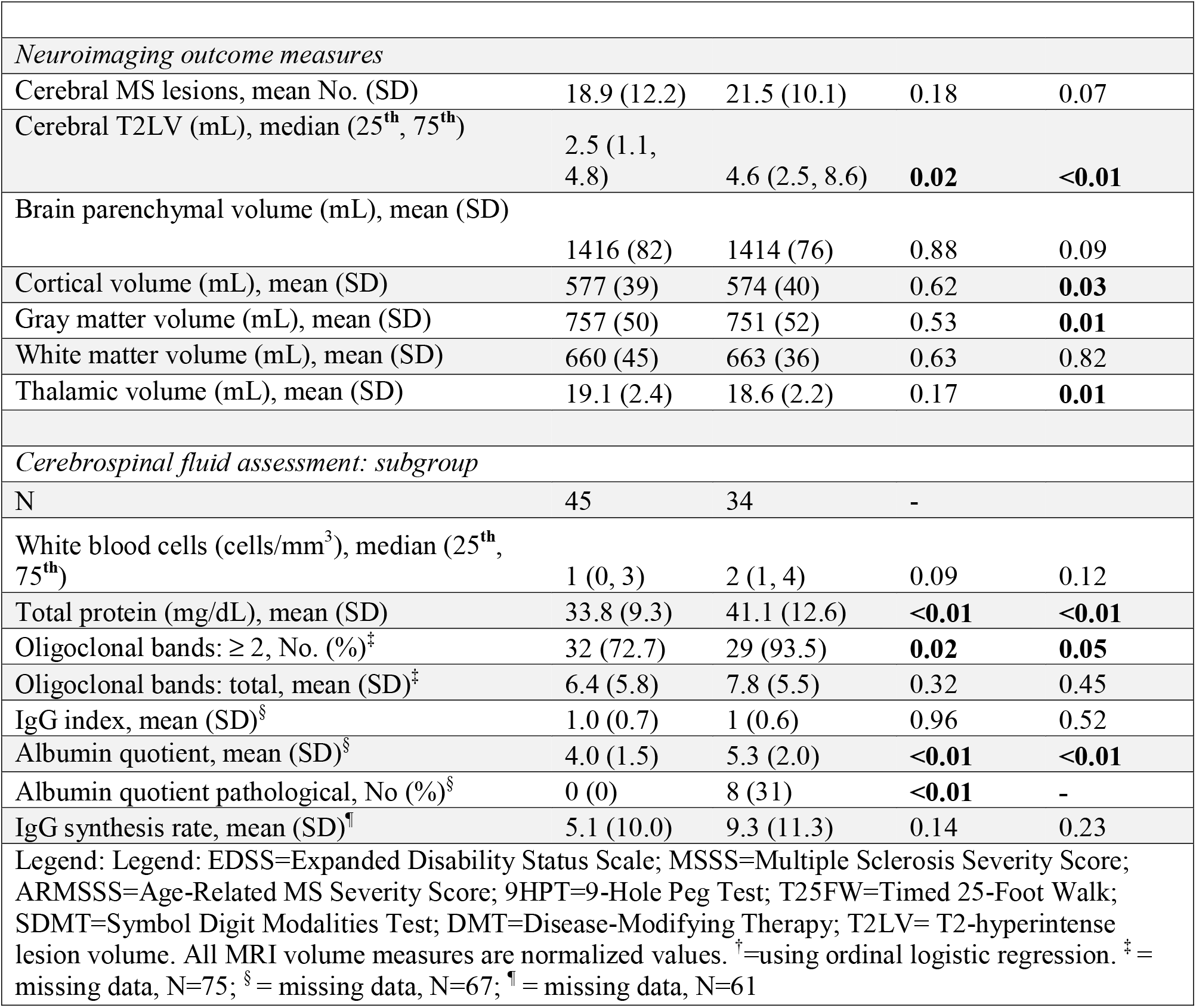
Clinical and demographic characteristics, neuroimaging, and CSF-associated outcomes stratified by PRL

### Paramagnetic Rim Lesion Assessment

Patients with at least 1 PRL (n=63) represented 43% of the total cohort. These patients were younger than patients without PRL (51±11 vs. 46±12 years, p=0.01). All PRL (100%) were hypointense on comparison T1-weighted gradient echo imaging. Examples of PRL can be seen in Figures 1 and 2. Summaries of the clinical, demographic, MRI, and cerebrospinal fluid (CSF) data are presented in Table 2 (stratified by PRL status, i.e., the presence of at least one PRL). Of particular note, PRL status was associated with shorter disease duration (14.5±10.1 vs. 10.5±9.1 years, p=0.01), greater disease severity (MSSS: 2.8±2.1 vs. 3.7±2.9, p=0.05), poorer upper extremity dexterity (13±3.7 vs. 14.4±4.2 seconds, p=0.05), higher likelihood of being on a more efficacious DMT (ordinal regression model, p=0.01), and larger T2LV (median 2.5 vs. 4.6 mL, p=0.02). CSF analysis showed associations between PRL status and higher total protein (33.8±9.3 vs. 41.1±12.6 mg/dL, p<0.01) and albumin quotient (4.0±1.5 vs. 5.3±2.0, p<0.01), as well as a higher likelihood of having CSF-specific oligoclonal bands (≥2; 72.7% vs. 93.5%, p=0.02). There was a trend toward a greater number of CSF leukocytes in patients with at least one PRL (median 1 vs 2 cells/mm^3^, p=0.09). After adjusting for age in a regression model with robust standard errors, these associations generally became stronger, with PRL status also being associated with decreased normalized cerebral brain matter volumes, most notably of cortex (adjusted p=0.03), total gray matter (adjusted p=0.01), and thalamus (adjusted p=0.01). A cognitive measure (the SDMT) was also associated with PRL after adjustment for age (adjusted p=0.02). Having a pathologically high (age-adjusted) albumin quotient was exclusively observed in patients with at least one PRL (p<0.01). PRL were also categorized into three groups (0, 1–3, ≥4 PRL) for comparison with prior studies [4, 14] (Table S1). After adjustment for age, a highly similar pattern of associations was seen compared to the dichotomous results.

**Figure 1:**
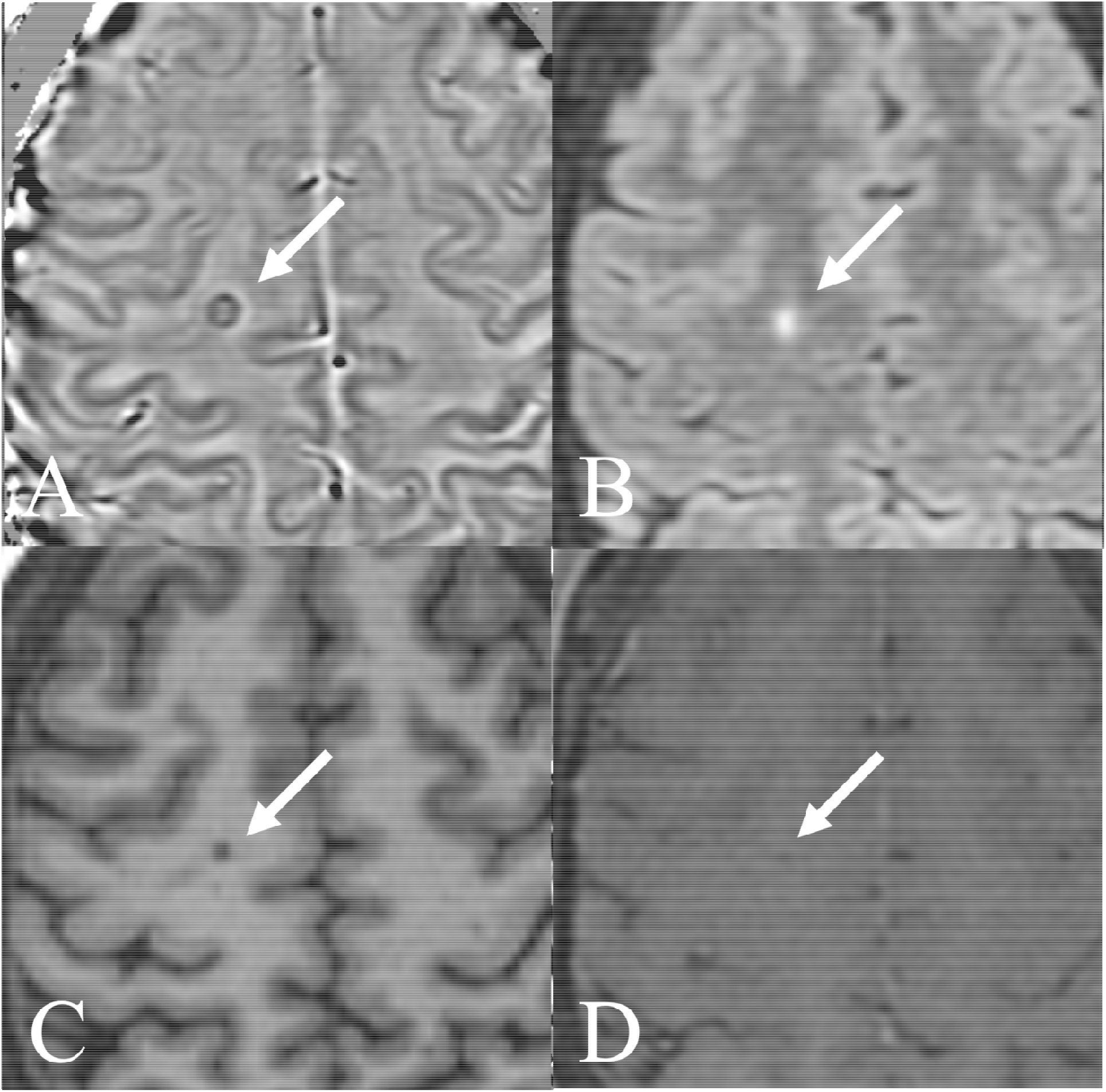
A woman in her 30s with relapsing MS showing a clear paramagnetic rim lesion as depicted on 3T MRI axial (A) filtered-phase T2* susceptibility image, (B) FLAIR, (C) T1-weighted GRE and (D) post-contrast T1-weighted spin-echo imaging.

**Figure 2:**
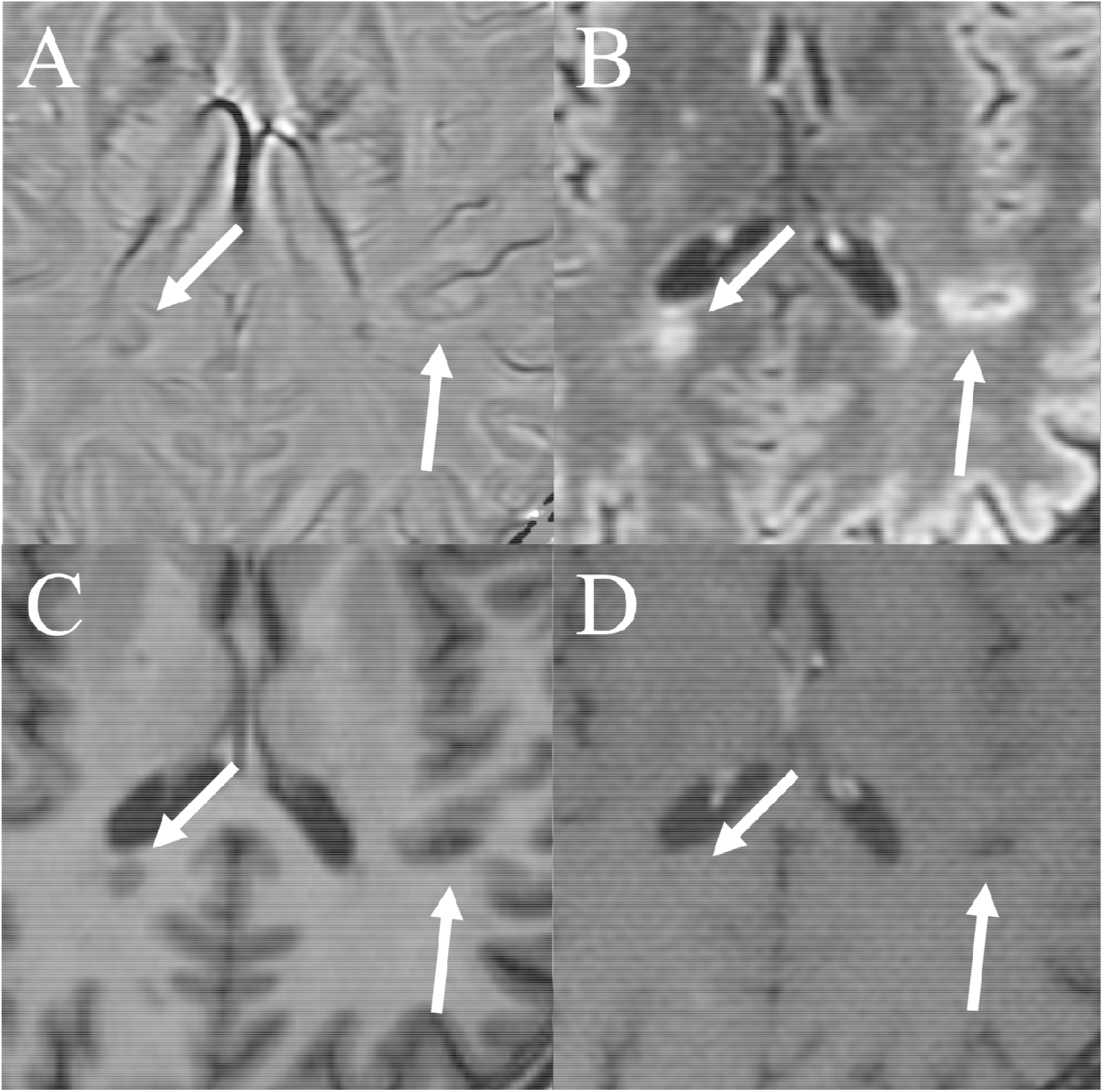
A man in his 30s with relapsing MS showing multiple paramagnetic rim lesions as depicted on 3T MRI axial (A) filtered-phase T2* susceptibility image, (B) FLAIR, (C) T1-weighted GRE and (D) post-contrast T1-weighted spin-echo imaging.

Given the possibility that clinical or MRI disease activity could mediate a confound between PRL and blood-brain-barrier permeability, we performed additional sensitivity analyses on subgroups of patients who: (1) had no evidence of clinical or MRI disease activity in the 3 months prior to the lumbar puncture (*n*=38), and (2) no gadolinium-enhancing lesions on an MRI performed within one month of the lumbar puncture (*n*=26) (Supplementary Tables S2 and S3). Notably, in both subgroup analyses there remained pathological associations with PRL status (≥1) and the albumin quotient, before and after adjusting for age (all p≤0.02). Importantly, there were also no significant differences in CSF protein-related measures between subgroups with and without disease activity (mean CSF protein 37.7±12.0 vs 36.0±11.4, p=0.50; mean CSF albumin quotient 4.6±1.9 vs 4.4±1.6, p=0.67; other CSF data not shown).

Last, following the observation of a significant association between PRL and T2LV, additional sensitivity analyses were performed to regress out this potential confounding factor. After adjustment, ≥1 PRL remained associated with higher CSF protein (p=0.01), pathologically high albumin quotient (p=0.02), and worse performance on the 9-hole peg test (p=0.04). These results are summarized in the supplemental materials (Table S4).

## Discussion

The paramagnetic (iron) rim lesion is an emerging MRI biomarker for the noninvasive assessment of chronic active lesions in MS. In this cross-sectional cohort study, we provide evidence for a novel association between PRL and the presence of greater blood-brain-barrier permeability and chronic intrathecal inflammation as measured by CSF protein/albumin quotient and the presence of oligoclonal bands, respectively. PRL are also associated with greater MS disease severity, worse upper extremity fine motor control, larger T2LV, and slower age-adjusted information processing along with greater age-adjusted cerebral atrophy. This last observation was particularly significant in the grey matter. These findings overall support the potential importance of PRL as biomarkers of MS disease severity and intrathecal inflammation that may be amenable to targeted therapies.

Our observations regarding the clinical and demographic associations with PRL are in general agreement with the literature. In multiple other studies that examined clinical or MRI associations with PRL, the authors universally conclude that PRL have pathogenic associations. A recent meta-analysis of 29 studies of PRL found a pooled prevalence of 40.6% of MS patients having at least one PRL using heterogeneous methods [16]; this is very close to our own finding of 43%. Most studies, including our own, have not found any association between PRL and sex; this is supported by the recent meta-analysis as well [16]. In the largest of these studies, an international, multicenter cohort of 329 patients was assessed using a 3D-EPI susceptibility protocol with an isotropic resolution of 0.65mm voxels. PRL were categorized into 3 groups (none, 1–3, or ≥4); the authors observed significant “dose-dependent” pathological associations between PRL and the EDSS score as well as the MSSS. Two other studies examined PRL at 3T (N=66) and 7T (N=100) and did not find baseline associations with EDSS but noted that PRL were a significant factor in predicting disability worsening over 2.9-year [5] and 1.4-year [17] follow-up. Few studies have examined associations with disease-modifying therapies; our data here agree with one other group’s that patients with PRL were treated with higher-efficacy therapies [15]. Given that treating clinicians did not clinically assess for PRL at our center at the time these data were assessed, this likely reflects a clinical interpretation of greater disease severity or prior treatment failure requiring a higher-efficacy treatment. Last, we observed an inverse association between PRL status and age; that is, ≥1 PRL is more likely to occur in younger individuals. Although several other studies have not observed any association with age [4, 15], our results agree with a recent meta-analysis of 29 studies of PRL [16], which concluded that PRL prevalence increased with younger age. Since PRL are more likely to form in lesions that develop in people older than 30 [18], the association with younger age seen here is probably driven by slow disappearance of PRL over time [18, 19].

Several previous studies have examined MRI associations of PRL, uniformly concluding that greater numbers of PRL are associated with higher T2LV [4, 5, 17, 20]. Fewer studies have examined, or been powered to determine, associations between PRL and quantified MRI volumes. Here, we find that after a statistical adjustment for age, our results indicate an association between PRL and more advanced gray matter atrophy; this finding is in alignment with data from Absinta et al. [4], who observed greater cerebral atrophy (N=192) in association with higher prevalence of PRL. After statistical adjustment for T2LV, PRL-cerebral atrophy associations were no longer significant; this implies that PRL may not have a large contribution to cross-sectional cerebral volume loss independent of total inflammatory lesion load.

This is the first study to our knowledge to formally characterize associations between PRL and the routine clinical CSF profile of patients. Overall, we observe a pathological association between PRL and CSF markers, with evidence for greater permeability of the blood-brain barrier, as suggested by a higher protein and albumin quotient, and higher likelihood of having oligoclonal bands. PRL were also observed to be exclusively associated with the age-adjusted “pathologically” high albumin quotients. These observations support existing evidence that PRL represent chronic active MS lesions that likely contribute to (focal) intrathecal inflammation, perhaps via the histologically confirmed presence of iron-rich, activated microglia at the lesion borders [3, 18]. Importantly, the correlations between PRL and elevated CSF protein/albumin quotient remain robust after statistical adjustment for T2LV, further suggesting that these findings are not related to generalized inflammatory disease burden. Oligoclonal bands were observed more frequently in patients with PRL, providing preliminary support for the proposal that PRL (combined with the central vein sign [21]) could act as diagnostic biomarkers in place of OCB, as recently explored in a cohort of CIS patients [22]. However, we did not find PRL to be associated with the magnitude of intrathecal immunoglobulin production as represented by the IgG index and synthesis rate; given the typically high correlation between elevated IgG index and OCB, this observation requires further investigation. Finally, an important consideration in the interpretation of the CSF results is the time lag between the association; here the lumbar puncture was performed on average 3.8 years prior to the time of the MRI scan. This methodological challenge introduces temporal uncertainty into the results, as well as the possibility of reverse causality—namely that a more permeable blood-brain barrier may predispose patients toward the development of PRL rather than the opposite. Future research will be addressing this question with longitudinal datasets and larger sample sizes.

Strengths of this study include a large cohort with detailed clinical and radiographic characterization; the use of an out-of-the-box susceptibility protocol to assess for PRL enhances the real-world translational value of our findings. Multiple sensitivity analyses support results. This is also the first study to our knowledge to examine CSF-based laboratory correlations of these lesions. Limitations include the retrospective, cross-sectional design, and use of a convenience cohort. This methodology could introduce selection bias; here the CSF cohort demonstrated shorter disease duration and greater disease severity, suggestive of a potential selection bias based on clinician determination of the need to perform a lumbar puncture. Much of this cohort also demonstrated clinical or radiological disease activity at the time of the lumbar puncture, which could have biased results. However, ameliorating this concern are prior evidence demonstrating the long-lasting nature of PRL (on the order of many years) [19, 23], as well as the consistent associations between PRL and CSF protein-related measures in subgroup analyses of patients without disease activity potentially contributing to CSF data. This study is additionally limited by having only a single rater assess images for the presence of PRL. It has been previously shown that inter-rater comparisons at 3T are substantial but imperfect, with kappa values around 0.7 [24]. Challenges related to PRL selection include the assessment of T2-FLAIR lesions with confluent characteristics (ill-defined borders), containing heterogeneous paramagnetic contents, and differentiating borders in the context of dense vascularity such as those encountered in the periventricular area [11]. Some paramagnetic rims also had a low signal-to-noise ratio (i.e. faint), highlighting the limitations of subjective interpretation. This limitation is somewhat mitigated by the categorical analysis demonstrating some “dose-dependent” effects such that more PRL are associated with worse clinical outcomes. Finally, patients had a wide variety of DMT histories (beyond just the treatment at the time of MRI), which were not evaluated here as it was beyond the scope of this study, and the combinatorics would reduce the effective sample size. Nonetheless, treatment may be an important factor in both the development and evolution of PRL; this remains an important area of future longitudinal research.

## Conclusions

The observation of at least one PRL connotes pathological importance in MS; PRL are also associated with CSF findings of greater blood-brain barrier permeability, supporting their characterization as noninvasive biomarkers of chronic intrathecal inflammation and endothelial dysfunction.

## Data Availability

All data produced in the present study are available upon reasonable request to the authors by qualified investigators.

