## Supplementary material for "Paramagnetic rim lesions are associated with pathogenic CSF profiles and worse clinical outcomes in multiple sclerosis: a retrospective cross-sectional study": Table S1

| Table S1: Clinical and demographic characteristics, neuroimaging, and CSF-associated outcomes stratified by PRL classified into groups | | | | |  |
| --- | --- | --- | --- | --- | --- |
|  | No Rims | 1–3 Rims | 4+ Rims | P-value, unadjusted | P-value, age-adjusted Sig. |
| *Demographic and Clinical Data* | | | | | |
| N (% total) | 84 (57.1) | 49 (33.3) | 14 (9.5) | - | - |
| Age, mean (SD), y | 51.1 (11.4) | 47.9 (11.4) | 38.8 (9.3) | **<0.01** | - |
| Sex, female, No. (%) | 68 (81.0) | 35 (71.4) | 10 (71.4) | 0.40 | 0.11 |
| Multiple sclerosis disease category | | | | | |
| - CIS/RR, No. (%) | 69 (82.1) | 41 (83.7) | 10 (71.4) | 0.57 | 0.11 |
| - SP/PP, No. (%) | 15 (17.9) | 8 (16.3) | 4 (28.6) |  |  |
| Disease duration, mean (SD), y | 14.5 (10.1) | 11.5 (9.6) | 7.1 (6.1) | **0.02** | 0.16 |
| EDSS score, median (25^th^, 75^th^) | 2.0 (1.5, 3.75) | 2.0 (1.5, 4.0) | 1.5 (1.0, 4.0) | 0.65 | 0.22 |
| MSSS, mean (SD) | 2.8 (2.1) | 3.6 (2.8) | 3.8 (3.1) | 0.14 | **0.04** |
| ARMSS score, mean (SD) | 3.2 (2.3) | 3.7 (2.6) | 4.3 (2.5) | 0.21 | 0.23 |
| Nine-hole peg test, mean (SD), sec | 13 (3.7) | 14.2 (4.3) | 15.1 (4.1) | 0.11 | **<0.01** |
| Timed 25-foot walk, mean (SD), sec | 6.2 (3.7) | 6.1 (2.0) | 7.3 (4.6) | 0.51 | 0.27 |
| SDMT score, mean (SD) | 53 (13.7) | 49.3 (14.4) | 48.5 (14) | 0.28 | **0.01** |
| Disease modifying therapy, No. (%) | | | |  |  |
| - Naïve | 6 (7.1) | 1 (2.0) | 0 (0.0) | **<0.01**^†^ | **0.02^†^** |
| - None at assessment | 20 (23.8) | 6 (12.2) | 3 (21.4) |  |  |
| - Low efficacy | 28 (33.3) | 18 (36.7) | 1 (7.1) |  |  |
| - Moderate efficacy | 7 (8.3) | 7 (14.3) | 1 (7.1) |  |  |
| - High efficacy | 23 (27.4) | 17 (34.7) | 9 (64.3) |  |  |
| *Neuroimaging outcome measures* | | | | | |
| Cerebral MS lesions, mean No. (SD) | 18.9 (12.2) | 21.7 (10.8) | 20.9 (7.9) | 0.39 | 0.07 |
| Cerebral T2LV (mL), median (25^th^, 75^th^) | 2.52 (1.08, 4.85) | 4.69 (2.46, 7.79) | 3.80 (2.56, 8.56) | **<0.01** | **<0.01** |
| Brain parenchymal volume (mL), mean (SD) | 1416.4 (81.7) | 1411.4 (77.7) | 1425 (69.7) | 0.84 | 0.06 |
| Cortical volume (mL), mean (SD) | 577.2 (38.9) | 570.5 (42.2) | 586 (31.7) | 0.38 | **0.04** |
| Gray matter volume (mL), mean (SD) | 756.5 (49.7) | 746.9 (52.9) | 766.3 (45.2) | 0.37 | **0.02** |
| White matter volume (mL), mean (SD) | 659.9 (44.7) | 664.5 (37.7) | 658.6 (31.6) | 0.80 | 0.43 |
| Thalamic volume (mL), mean (SD) | 19.1 (2.4) | 18.5 (2.2) | 18.7 (2) | 0.38 | **<0.01** |
| *Cerebrospinal fluid assessment: subgroup* | | | | | |
| N | 45 | 28 | 6 | - |  |
| White blood cells (cells/mm^3^), median (25^th^, 75^th^) | 1.8 (2.1) | 4.1 (10.4) | 8.3 (12.5) | 0.08 | 0.08 |
| Total protein (mg/dL), mean (SD) | 33.8 (9.3) | 41.7 (13.2) | 38.2 (9.9) | **0.01** | **<0.01** |
| Oligoclonal bands: ≥ 2, No. (%)^‡^ | 32 (72.7) | 23 (92) | 6 (100) | 0.07 | **0.01** |
| Oligoclonal bands: total, mean (SD)^‡^ | 6.4 (5.8) | 6.7 (4.6) | 12.2 (7.2) | 0.06 | 0.20 |
| IgG index, mean (SD)^§^ | 1.0 (0.7) | 0.9 (0.4) | 1.2 (0.9) | 0.48 | 0.76 |
| Albumin quotient, mean (SD)^§^ | 4.0 (1.5) | 5.6 (2.0) | 4.2 (1.6) | **<0.01** | **0.02** |
| Albumin quotient pathological, No (%)^§^ | 0 (0) | 7 (35) | 1 (20) | **<0.01** | **-** |
| IgG synthesis rate, mean (SD)^¶^ | 5.2 (10.1) | 8.1 (10.9) | 12.9 (15.1) | 0.31 | 0.28 |
| Legend: EDSS=Expanded Disability Status Scale; MSSS=Multiple Sclerosis Severity Score; 9HPT=9-Hole Peg Test; T25FW=Timed 25-Foot Walk; SDMT=Symbol Digit Modalities Test; DMT=Disease-Modifying Therapy; T2LV= T2-hyperintense lesion volume. All MRI volume measures are normalized values. ^†^=using ordinal logistic regression; ^‡^ = missing data, N=75; ^§^ = missing data, N=67; ^¶^ = missing data, N=61 | | | | | |

| Table S2: PRL-CSF associations, subgroup analysis of patients without prior clinical or MRI disease activity in the 3 months prior to lumbar puncture | | | | |
| --- | --- | --- | --- | --- |
|  | No PRL | ≥1 PRL | P-value, unadjusted | P-value, age-adjusted Sig. |
| *Cerebrospinal fluid assessment: subgroup* |  |  |  |  |
| N | 27 | 16 | - |  |
| White blood cells (cells/mm^3^), median (25^th^, 75^th^) | 1 (1, 3) | 1 (1, 1.5) | 0.70 | 0.36 |
| Total protein (mg/dL), mean (SD) | 35.0 (8.9) (9.3) | 42.4 (15.2) | **0.05** | **0.04** |
| Oligoclonal bands: ≥ 2, No. (%)^‡^ | 16 (62) | 14 (93) | **0.03** | **0.01** |
| Oligoclonal bands: total, mean (SD)^‡^ | 6.2 (6.4) | 6.1 (4.6) | 0.96 | 0.70 |
| IgG index, mean (SD)^§^ | 1.0 (0.7) | 1.1 (0.8) | 0.74 | 0.95 |
| Albumin quotient, mean (SD)^§^ | 4.1 (1.4) | 5.5 (2.4) | **0.02** | **0.02** |
| Albumin quotient pathological, No (%)^§^ | 0 (0) | 5 (38) | **<0.01** | **-** |
| IgG synthesis rate, mean (SD)^¶^ | 5.3 (10.0) | 10.3 (14.4) | 0.24 | 0.35 |
| Legend: ^‡^ = missing data, N=43; ^§^ = missing data, N=38; ^¶^ = missing data, N=34 | | | | |

| Table S3: PRL-CSF associations, subgroup analysis of patients with no gadolinium enhancing lesions on MRI within 1 month of lumbar puncture | | | | |
| --- | --- | --- | --- | --- |
|  | No PRL | ≥1 PRL | P-value, unadjusted | P-value, age-adjusted Sig. |
| *Cerebrospinal fluid assessment: subgroup* |  |  |  |  |
| N | 20 | 11 | - |  |
| White blood cells (cells/mm^3^), median (25^th^, 75^th^) | 1 (1, 3) | 1 (1, 4) | 0.83 | 0.55 |
| Total protein (mg/dL), mean (SD) | 34.4 (7.2) | 41.7 (14.1) | 0.06 | 0.10 |
| Oligoclonal bands: ≥ 2, No. (%)^‡^ | 13 (65) | 9 (90) | **0.03** | 0.11 |
| Oligoclonal bands: total, mean (SD)^‡^ | 6.5 (6.4) | 6.4 (4.5) | 0.97 | 0.95 |
| IgG index, mean (SD)^§^ | 1.0 (0.7) | 0.9 (0.5) | 0.80 | 0.76 |
| Albumin quotient, mean (SD)^‖^ | 4.0 (1.2) | 5.7 (1.9) | **0.01** | **0.02** |
| Albumin quotient pathological, No (%) | 0 (0) | 3 (33) | **<0.01** | **-** |
| IgG synthesis rate, mean (SD)^¶^ | 6.2 (11.0) | 8.6 (14.2) | 0.63 | 0.66 |
| Legend: ^‡^ = missing data, N=30; ^§^ = missing data, N=27; ^‖^ = missing data, N= 26; ^¶^ = missing data, N=25 | | | | |

| Table S4: Linear regression model of the association between PRL and clinical, MRI, and CSF measures adjusting for the volume of T2-hyperintense lesions | | | | | | | |
| --- | --- | --- | --- | --- | --- | --- | --- |
|  | Age- and T2LV-adjusted associations with dichotomous PRL (≥1) | | |  | Age- and T2LV-adjusted associations with categorical PRL (0, 1-3, 4+) | | |
|  | *β* | 95% CI | *p* |  | *β* | 95% CI | *p* |
| Age (yrs) | - | - | **-** |  | - | - | - |
| **Sex** |  |  |  |  |  |  |  |
| - Male | - | - | - |  |  |  |  |
| - Female | -0.13 | -0.27,0.02 | 0.08 |  | -0.1 | -0.21, 0.01 | 0.08 |
| MS disease phenotype |  |  |  |  |  |  |  |
| - RR/CIS | - | - | - |  |  |  |  |
| - PP/SP | 0.02 | -0.10,0.15 | 0.72 |  | 0.05 | -0.05, 0.15 | 0.32 |
| Disease duration | -2.94 | -5.55,-0.33 | **0.03** |  | -2.26 | -4.17,-0.35 | **0.02** |
| EDSS | 0.14 | -0.48,0.76 | 0.66 |  | 0.13 | -0.33,0.59 | 0.59 |
| MSSS | 0.9 | -0.07,1.87 | 0.07 |  | 0.69 | -0.07,1.44 | 0.08 |
| 9HPT | 1.21 | -0.07,2.49 | 0.06 |  | 1.19 | 0.19,2.19 | **0.02** |
| T25FW | 0.09 | -0.85,1.04 | 0.84 |  | 0.37 | -0.56,1.29 | 0.44 |
| SDMT | -2.64 | -7.14,1.86 | 0.25 |  | -2.33 | -5.86,1.20 | 0.19 |
| DMT efficacy | 0.38 | -0.05,0.81 | 0.08 |  | 0.33 | -0.01,0.67 | 0.06 |
| *Neuroimaging outcome measures* | | | | | | | |
| Cerebral MS lesions, No. | -5.47 | -27.02,16.07 | 0.62 |  | 0.59 | -2.00,3.18 | 0.65 |
| Cerebral T2LV (mL) | -7.97 | -19.58,3.64 | 0.18 |  | - | - | **-** |
| Brain parenchymal volume (mL) | -10.59 | -24.76,3.57 | 0.14 |  | -5.61 | -22.46,11.24 | 0.51 |
| Cortical volume (mL) | 5.12 | -7.11,17.34 | 0.41 |  | -5.2 | -13.97,3.58 | 0.24 |
| Gray matter volume (mL) | -0.26 | -0.77,0.24 | 0.30 |  | -7.03 | -18.33,4.27 | 0.22 |
| White matter volume (mL) | -5.47 | -27.02,16.07 | 0.62 |  | 1.42 | -7.31,10.14 | 0.75 |
| Thalamic volume (mL) | -7.97 | -19.58,3.64 | 0.18 |  | -0.22 | -0.62,0.18 | 0.28 |
| *Cerebrospinal fluid assessment: subgroup* | | | | | | | |
| White blood cells (cells/mm^3^) | 3.94 | -1.07,8.95 | 0.12 |  | 3.9 | -0.22,8.01 | 0.06 |
| Total protein (mg/dL) | 8.89 | 3.35,14.42 | **<0.01** |  | 6.36 | 1.81,10.90 | **0.01** |
| Oligoclonal bands: ≥ 2^‡^ | 0.17 | -0.02,0.36 | 0.07 |  | 0.13 | -0.00,0.26 | **0.05** |
| Oligoclonal bands: total^‡^ | 1.49 | -1.26,4.24 | 0.28 |  | 2.09 | -0.31,4.50 | 0.09 |
| IgG index^§^ | -0.16 | -0.58,0.26 | 0.44 |  | -0.1 | -0.51,0.31 | 0.63 |
| Albumin quotient^‖^ | 1.47 | 0.53,2.41 | **<0.01** |  | 0.87 | 0.01,1.73 | **0.05** |
| Abnormal albumin quotient^‖^ | 0.22 | 0.07,0.36 | **<0.01** |  | 0.14 | 0.02,0.26 | **0.03** |
| IgG synthesis rate^¶^ | 2.02 | -6.13,10.18 | 0.62 |  | 1.63 | -5.77,9.04 | 0.66 |
| Legend: EDSS=Expanded Disability Status Scale; MSSS=Multiple Sclerosis Severity Score; 9HPT=9-Hole Peg Test; T25FW=Timed 25-Foot Walk; SDMT=Symbol Digit Modalities Test; DMT=Disease-Modifying Therapy; T2LV= T2-hyperintense lesion volume. All MRI volume measures are normalized values. ^†^=using ordinal logistic regression; ^‡^ = missing data, N=75; ^§^ = missing data, N=67; ^‖^ = missing data, N= 65; ^¶^ = missing data, N=59 | | | | | | | |
